# Airway Epithelium Respiratory Illnesses and Allergy (AERIAL) birth cohort: study protocol

**DOI:** 10.1101/2023.04.29.23289314

**Authors:** Elizabeth Kicic-Starcevich, David G Hancock, Thomas Iosifidis, Patricia Agudelo-Romero, Jose A Caparros-Martin, Desiree Silva, Lidija Turkovic, Peter N Le Souef, Anthony Bosco, David J Martino, Anthony Kicic, Susan L Prescott, Stephen M Stick

## Abstract

**Introduction:** Recurrent wheezing disorders including asthma are complex and heterogeneous diseases that affect up to 30% of all children, contributing to a major burden on children, their families, and global healthcare systems. It is now recognized that a dysfunctional airway epithelium plays a central role in the pathogenesis of recurrent wheeze, although the underlying mechanisms are still not fully understood. This prospective birth cohort aims to bridge this knowledge gap by investigating the influence of intrinsic epithelial dysfunction on the risk for developing respiratory disorders and the modulation of this risk by maternal morbidities, *in utero* exposures, and respiratory exposures in the first year of life.

**Methods and Analysis:** The Airway Epithelium Respiratory Illnesses and Allergy (AERIAL) study is nested within the ORIGINS Project and will monitor 400 infants from birth to five years. The primary outcome of the AERIAL study will be the identification of epithelial endotypes and exposure variables that influence the development of recurrent wheezing, asthma, and allergic sensitisation. Nasal respiratory epithelium at birth to six weeks, one, three, and five years will be analysed by bulk RNA-seq and DNA methylation sequencing. Maternal morbidities and *in utero* exposures will be identified on maternal history and their effects measured through transcriptomic and epigenetic analyses of the amnion and newborn epithelium. Exposures within the first year of life will be identified based on infant medical history as well as on background and symptomatic nasal sampling for viral PCR and microbiome analysis. Daily temperatures and symptoms recorded in a study-specific Smartphone App will be used to identify symptomatic respiratory illnesses.

**Ethics and Dissemination:** Ethical approval has been obtained from Ramsey Health Care HREC WA-SA (#1908). Results will be disseminated through open-access peer-reviewed manuscripts, conference presentations, and through different media channels to consumers, ORIGINS families, and the wider community.

## INTRODUCTION

Recurrent wheezing disorders in children are extremely common and impose significant burdens on children, families, and communities, with 20-30% of all children developing recurrent wheezing and 10% receiving an asthma diagnosis [1,2]. These early-life disorders also have a significant impact on healthcare systems worldwide, representing one of the most common reasons for hospital presentation and contributing billions of dollars to direct and indirect economic and healthcare costs [2,3]. While significant advances have been made in our understanding of disease pathophysiology, endotypes, and triggers, there are still major knowledge gaps leading to an ongoing lack of optimal diagnostic, disease prevention, and treatment options [4,5].

The pathogenesis of recurrent wheezing represents a complex interplay between intrinsic genetic, cellular, and structural factors, which are then modulated by external pathogen and environmental exposures. While deficient immune responses and interferon signalling are well characterised in these conditions [6], the airway epithelium has been increasingly recognised as a key contributor to this disease process [7], but the exact mechanisms are still unclear. As the first contact point for inhaled pathogens and particles, the airway epithelium plays an essential role in barrier function, mucociliary clearance, and immune/inflammatory responses in the airway [8]. To interrogate disease pathogenesis and susceptibility mechanisms in the airway epithelium, obtaining airway tissues is essential, but limited by ease of access [9]. While there exists differences in epithelial cell composition and gene expression between nasal, tracheal, and bronchial regions [10], strongly conserved gene signatures between nasal and tracheal samples have been described in both health and disease [9], supporting the nose as a minimally invasive site for interrogating global epithelial function throughout the airway. Primary airway epithelial cells isolated from both the upper and lower airways of young children with asthma have been shown to exhibit essential functional characteristics with compromised barrier function [11,12], wound repair [13-16], and innate immune responses [17], referred to as the “vulnerable epithelium” [18]. These observations of an intrinsically impaired epithelium are supported by genome-wide associations studies that have highlighted several genes variants that are expressed in epithelial cells as risk factors for asthma development [19,20].

Several key variables have been identified that further modulate this intrinsic epithelial risk. Maternal morbidities such as asthma, have been shown to be associated with an increased risk for childhood asthma development [21]. *In utero* exposures have also been linked to the development of recurrent wheezing/asthma in early-life including infection, allergen, medication/antibiotic, and pollutant exposures, with maternal smoking typically showing the strongest association [22-24]. The amnion provides a readily available, non-invasive tissue source to detect and measure the influence of maternal morbidities and *in utero* exposures after birth [25-27]. In support of this, transcriptional and epigenetic changes in the placenta and amnion have been described following *in utero* exposures such as infection or smoking [25-27]. The amnion is also a source of epithelial cells that have been exposed to similar factors as the developing respiratory epithelium of the foetus/newborn [25-27]. In addition to maternal morbidities and *in utero exposures*, early-life exposures to respiratory viruses and bacteria, as well as environmental allergens, pollutants, and cigarette smoke have all shown an association with subsequent recurrent wheezing and asthma development [28-31]. One mechanism through which exposures appear to modulate future respiratory outcomes is through epigenetic reprogramming of host cells, with distinct methylation profiles being observed in respiratory epithelial cells from healthy, atopic, and asthmatic children [32].

While published studies have focused on individual aspects of these disease mechanisms, there remains a need for more integrated studies investigating multiple factors both in combination and longitudinally. We have therefore designed this prospective longitudinal birth cohort study as an integrated platform to interrogate key factors influencing the development of recurrent wheezing, asthma, and allergic sensitisation across the first 5 years of life. We hypothesise that intrinsic epithelial vulnerability is a precursor to adverse respiratory outcomes in childhood, with this predisposition being further modulated by maternal morbidities, *in utero* exposures, and exposures within the first year of life.

The specific aims of this cohort study are:

1. To establish whether the vulnerable respiratory epithelium is detectable at birth, epigenetically regulated, and can be used to predict respiratory outcomes within the first 5 years of life.
2. To determine whether maternal morbidities and *in utero* exposures influence the development of the vulnerable epithelium at birth.
3. To determine whether the amnion can be used as a surrogate marker for antenatal exposures and/or vulnerable respiratory epithelium of the newborn.
4. To determine the role of the microbiome and respiratory virus exposures within the first year of life on the development of respiratory outcomes in those with vulnerable epithelium.

## METHODS AND ANALYSIS

### Study Design and Setting

The Airway Epithelium Respiratory Illnesses and Allergy (AERIAL) study is nested within the ORIGINS Project, a large Western Australian longitudinal, prospective birth cohort study, following children from the first trimester of their mother’s pregnancy through to five years of age [33]. Participant families for the ORIGINS Project and AERIAL study are recruited at Joondalup Health Campus, a public/private partnership hospital servicing a culturally and socioeconomically diverse region in Perth, Western Australia [33]. Recruitment into the AERIAL Study commenced in August 2020 during the COVID-19 pandemic and associated public health restrictions in Western Australia. The study is required to adhere to the guidelines set by the Western Australia Government and Health Department for participant interactions and sample collection.

### Participants and Recruitment

The AERIAL study will recruit 400 families from the ORIGINS Project, during a routine antenatal visit. Mothers will be consented into the study prior to delivery, while newborns will be consented as active participants after birth. Inclusion criteria for the AERIAL study is sufficient understanding of written/spoken English to complete consent/study procedures and access to a smartphone that can utilise the study App. To maintain diversity in the cohort, the only exclusions for this study are participant births with a gestational age less than 32 weeks, any significant genetic anomalies, and any significant perinatal complications.

### Participant and Consumer Involvement

AERIAL is an interactive longitudinal study which requires a high level of participation from the families, particularly within the first year of their child’s life, with daily temperature checks, recording of symptoms, and collection of four background nasal/throat swabs (three, six, nine, twelve months), as well as repeated symptomatic swabs during illness. To ensure high data and sample integrity, the study consulted extensively with consumers in two consumer reference groups (one which included ORIGINS participants) to develop the protocol which prioritised reduced burden as well as maximised adherence and retention. All consumers insisted on simplicity and ease of monitoring, communication, and daily recordings. This led to the development of a Smartphone app AERIAL TempTracker App enabling real time viral detection in the AERIAL cohort.

### Outcome measures

The primary outcome of the AERIAL study will be the characterisation of vulnerable epithelial endotype(s) and exposure variables that influence the development of recurrent wheezing, asthma, and allergic sensitisation at one, three, and five years of age. The presence of these endpoints will be determined independently of downstream analyses and will be based on serial review of each participants medical history during both AERIAL and ORIGINS study visits. Recurrent wheezing (at one, three, and five years of age) will be defined as the presence of two or more episodes of parent-reported or medically documented wheezing [34]. Asthma (at three and five years) will defined based on a diagnosis made by the participant’s treating physician or according to national guidelines [35] as chronic signs and symptoms suggestive of asthma with a documented treatment response. Allergic sensitisation will be defined as a positive skin-prick test result (at one, three, and five years as part of their ORIGINS Project clinic visit [33]), according to Australasian Society of clinical Immunology and Allergy guidelines [36].

Secondary outcomes include:

1. Investigating the use of the amnion as a surrogate marker for a child’s respiratory epithelium at birth and as a tool for classifying the epigenetic/transcriptomic effects of maternal morbidities and *in utero* exposures at birth.
2. Characterising the influence of epithelial endotypes and symptomatic viral infections on the development and maintenance of the microbiome in early life.
3. Identifying microbial signatures associated with protection or susceptibility to viral infections.

### Data Collection, Processing, and Analyses

The samples and data to be collected at each timepoint are presented in Figure 1 and Table 1.

**Figure 1.**
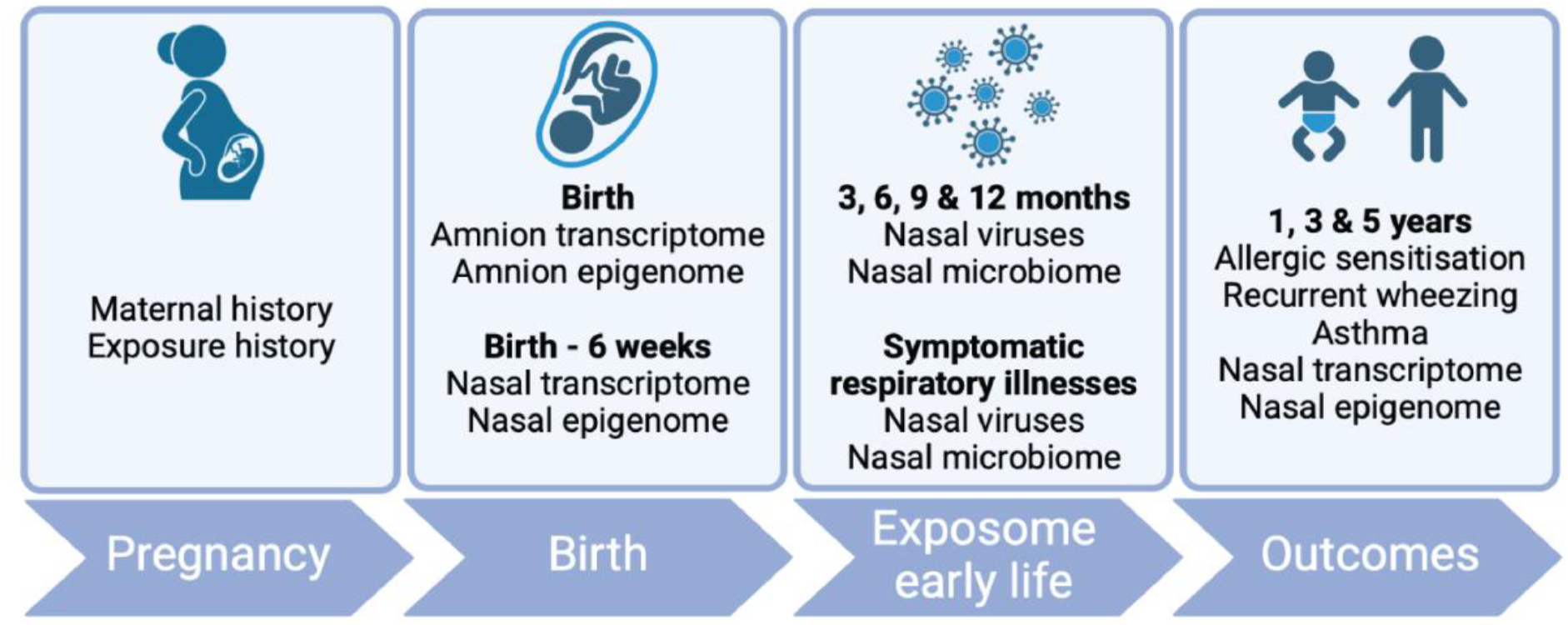
AERIAL Study Schema. Produced using BioRender.com

**Table 1:** AERIAL Study schedule of assessments.

|  | Timepoints |  |  |  |  |  |  |  |  |
| --- | --- | --- | --- | --- | --- | --- | --- | --- | --- |
|  | Antenatal | Newborn | Background |  |  | Symptomatic Illnesses | Follow-Up |  |  |
|  |  | Birth – 6 wks | 3mo | 6m | 9mo | < 1yr | 1yr | 3yr | 5yr |
| **Maternal Variables** |  |  |  |  |  |  |  |  |  |
| Socioeconomic/Demographic | X |  |  |  |  |  |  |  |  |
| Medical History | X |  |  |  |  |  |  |  |  |
| Smoking | X |  |  |  |  |  |  |  |  |
| Pregnancy History | X |  |  |  |  |  |  |  |  |
| Medications/Vaccinations | X |  |  |  |  |  |  |  |  |
| **Infant Variables** |  |  |  |  |  |  |  |  |  |
| Demographics |  | X |  |  |  |  |  |  |  |
| Birth History |  | X |  |  |  |  |  |  |  |
| Growth/Development |  | X | X | X | X |  | X | X | X |
| Medical History |  | X | X | X | X | X | X | X | X |
| **Amnion** |  |  |  |  |  |  |  |  |  |
| Epigenome |  | X |  |  |  |  |  |  |  |
| Transcriptome |  | X |  |  |  |  |  |  |  |
| **Nasal Epithelial Brushings** |  |  |  |  |  |  |  |  |  |
| Epigenome |  | X |  |  |  |  | X | X | X |
| Transcriptome |  | X |  |  |  |  | X | X | X |
| **Nasal Pathogen Swabs** |  |  |  |  |  |  |  |  |  |
| Viral Swab |  |  | X | X | X | X |  |  |  |
| Microbiome |  |  | X | X | X | X |  |  |  |
| **Endpoints** |  |  |  |  |  |  |  |  |  |
| Recurrent Wheezing |  |  |  |  |  |  | X | X | X |
| Asthma |  |  |  |  |  |  | X | X | X |
| Allergic Sensitisation |  |  |  |  |  |  | X | X | X |

#### Clinical Data Collection

Participant clinical data used in the AERIAL Study will be collected as part of the ORIGINS Project[33] and includes:

1. Maternal demographics, medical history (including asthma/atopy), *in utero* exposures (including smoking exposure), and pregnancy data.
2. Infant demographics, medical history, body composition, and growth/developmental assessments.

The AERIAL study will also collect additional information on respiratory illness, wheezing, allergies, and relevant medication use (including asthma relievers/preventers) at scheduled study visits as well as at symptomatic illness events.

#### Symptomatic Respiratory Illnesses

The AERIAL study aims to capture all respiratory illnesses within the first year of life. Parents will measure daily temperatures using an infrared forehead thermometer and record symptoms of respiratory illness in a designed-for-purpose Smartphone App, the AERIAL TempTracker App (Telethon Kids Institute). Symptomatic respiratory illnesses requiring a nasal swab will be defined as a temperature >37.5°C and the presence of at least one respiratory symptom (runny/blocked nose, wet/dry cough, sneeze, headache, myalgia, chills, rigours, tiredness, sore throat, dyspnoea, loss of taste/smell, poor feeding, diarrhoea) OR temperature <37.5°C and the presence of at least two respiratory symptoms. The AERIAL App will also be used to monitor and encourage participant compliance with study procedures.

#### Amnion Sampling

Amnion for transcriptomic and epigenetic analyses will be collected from consenting mothers. After delivery, placentas will be examined by the delivery personnel according to Joondalup Health Campus standard operating procedures, placed in plastic bags, and stored at 4°C for transport to Telethon Kids Institute (Perth). Biopsy samples from each amniotic membrane will be taken within 72hours of delivery, snap frozen, and stored at -80°C until batch processing.

#### Nasal Epithelial Sampling

Nasal epithelial sampling for transcriptomic and epigenetic analyses will be performed at birth to six weekshome visit), and at one, three, and five years of age (clinic visits), by a trained study team member. For newborns, each nostril will be sampled using a Dent-O-Care 620 brush (Dent-o-Care, United Kingdom), as previously described [37]. and placed into a FluidX™tube (Azenta Life Sciences, USA) containing DNA/RNA Shield™ (Zymo research, USA). For one-, three-, and five-year nasal sampling, only one nostril will be used. All collected samples will be store at 4°C for transport and then stored at -80°C until batch RNA and DNA extraction.

#### Nasal Swabs for Virus and Microbiome Analyses

During the first year of life, nose/throat swabs for respiratory virus identification and microbiome analysis will be collected from separate nostrils during symptomatic respiratory illnesses (as described above) and at quarterly background checks (three, six, nine, and twelve months). Samples were initially to be taken by study staff at home visits; however, the protocol was adjusted to enable parent collection of swabs during community COVID-19 transmission to adhere to public health restrictions and ensure staff and participant safety.

For respiratory virus identification, the throat and one nostril [38] will be sampled with a swab (FLoQSwabs, Copan Group, Italy),placed into viral testing medium (eSwab, Copan Group, Italy), and stored at 4°C until transport to the commercial laboratory (Western Diagnostics Pathology) for testing of 8 common respiratory viruses (influenza ‘A’, influenza ‘B’, respiratory syncytial virus, human metapneumovirus, parainfluenza, rhinovirus, adenovirus, SARS-CoV-2) by multiplex PCR. For microbiome analyses, the other nostril will be sampled, the swab placed into DNA/RNA stabilisation medium (eNAT, Copan Group, Italy), transported at 4°C, and frozen at -80°C until batch processing.

#### Additional Biological Samples

Additional samples including maternal and infant blood, urine, saliva, and stool samples are collected at various timepoints and biobanked as part of the ORIGINS Project[33] and are available for secondary analysis as required during the AERIAL study. Participants of both the AERIAL and ORIGINS Project will be asked to consent to allow these secondary analyses on bio-banked samples.

#### Transcriptomic Profiling

RNA from amnion and nasal epithelial samples will be extracted using Zymo Quick-RNA Microprep Kits (Zymo Research, USA), according to manufacturer’s instructions, and stored at -80°C. Total RNA quantity, quality and integrity will be assessed by Qubit (ThermoFisher, USA) and the Agilent TapeStation (Agilent Technologies, USA). RNA samples will be sent on dry ice to a genomics core facility for whole-transcriptome library preparation and sequencing.

#### Epigenetic Profiling

For epigenetic analysis, DNA from amnion and nasal epithelial samples will be extracted using Chemagic DNA Blood Kits (PerkinElmer, USA), according to the manufacturer’s instructions, and stored at -80°C until analysis in bulk. DNA quantity will be assessed by Qubit (I ThermoFisher, USA). Capture DNA methylation sequencing (methyl-seq) using enzymatic conversion and target enrichment will be conducted at a genomics core facility using the TWIST Human Methylome Panel, covering more than 3.2 million CpGs.

#### Microbiome Profiling

Microbial DNA will be extracted using QIAamp DNA kit (QIAGEN, location). Positive extraction controls in the form of spike-in internal control samples and mock communities from ZymoBIOMICS (Zymo Research, USA) will be included. Bacterial load in DNA extracts will be quantified using TaqMan qPCR (ThermoFisher, USA)[39]. The bacterial biota will be profiled at strain level resolution by amplifying and sequencing the full-length *16S rRNA* gene.

### Statistical Analysis

#### Analytical Methods

Analyses will be performed on longitudinally acquired data, using generalised linear mixed effects models with random subject effects and best fitting covariance structure. Primary endpoints of recurrent wheezing, asthma, and allergic sensitisation will be assessed as binary outcomes (yes/no). Number of wheezing episodes will also be assessed as a discrete outcome. All models will be adjusted for interactions and confounders as required (including demographic and socioeconomic variables). Transcriptomic, methylation, and microbiome data will be pre-processed and analysed using our established in-house protocols and pipelines [9,39,40].

#### Sample Size

A total of 400 participants will be recruited into AERIAL to be followed for 5 years and will be randomly divided into discovery (300 participants) and validation (100 participants) cohorts for analysis. Sample size calculations are based on our published RNA-Seq data [9]. R package RNASeqPower [41] was used to determine sample sizes for the discovery cohort with an estimated outcome prevalence of 20%, library depth of 15 million, biological coefficient of variation of 0.5, power of 80%, a = 0.05, and a range of fold difference effect sizes. We will recruit 300 children as a discovery cohort to ensure retention of 260-270 children with adequate diary information and viral ascertainment of ~ 65% [42,43]. The validation cohort will be used to assess the ability of target biomarkers from the discovery cohort to predict the main respiratory outcomes of interest, using penalised logistic regression and receiver operating characteristic curves (AUC). 100 children will give us an approximate power of 0.95 (a = 0.05, R package pROC [44]) to detect a difference in AUC between 0.75 and 0.5 for recurrent wheezing, asthma, and atopic sensitisation endpoints.

## Data Availability

All data produced in the present work are contained in the manuscript

## ETHICS AND DISSEMINATION

The study is being conducted in accordance with the Helsinki Declaration and was approved by the Ramsey Health Care HREC WA-SA (#1908). Participating families may withdraw consent for the study at any time and privacy and confidentiality will be provided by assigning a unique AERIAL study ID at the time of consent. All specimens, reports, and data collected by AERIAL are identified by ID number only.

The de-identified results of the analysis and data will be made available and disseminated through open-access peer-reviewed manuscripts, local, national and international conference presentations, and through different media channels to the consumers, AERIAL/ORIGINS families, and the wider community.

## SUMMARY

The Airway Epithelium Respiratory Illnesses and Allergy study (AERIAL) will longitudinally characterise the determinates of recurrent wheezing, asthma, and allergic sensitisation in early life. While individual aspects of these disease processes have been interrogated previously, the key strengths of the AERIAL cohort are its longitudinal design, multi-omics approach, and robust collation of critical exposures both *in utero* and through the first year. The use of a Smartphone App will allow for reduced burden on participant families and study coordinators and support adjustments to evolving pandemic restrictions. Through an improved understanding of both intrinsic epithelial vulnerability and the external factors modulating its phenotypic manifestation, we hope to provide a foundation for new insights into disease pathogenesis unlocking novel opportunities for diagnostic and therapeutic development.

## AUTHOR CONTRIBUTIONS

Study concept and design: SS, PLS, DM, AB, AK, SP

Study design: LKS, PAR, JCM, TI, DH, LT, DS

Drafting of the manuscript: LKS, DH

Study management: LKS.

All authors read, critically reviewed, and approved the final manuscript.

## ACKNOWLEDGEMENTS

We would like to thank the contribution of the AERIAL families and the dedicated research team, for the recruitment, liaising and sample collection over the duration of the study.

This study is a sub-project of The ORIGINS Project. This unique long-term study, a collaboration between Telethon Kids Institute and Joondalup Health Campus, is one of the most comprehensive studies of pregnant women and their families in Australia to date, recruiting 10,000 families over a decade from the Joondalup and Wanneroo communities of Western Australia. We are grateful to all the ORIGINS families who support the project.

We would also like to acknowledge and thank the following teams and individuals who have made The ORIGINS Project possible: The ORIGINS Project team; Joondalup Health Campus (JHC); members of ORIGINS Community Reference and Participant Reference Groups; Research Interest Groups and the ORIGINS Scientific Committee; Telethon Kids Institute; City of Wanneroo; City of Joondalup; and Professor Fiona Stanley.

The ORIGINS Project has received core funding support from the Telethon Perth Children’s Hospital Research Fund, Joondalup Health Campus, the Paul Ramsay Foundation and the Commonwealth Government of Australia through the Channel 7 Telethon Trust. Substantial in-kind support has been provided by Telethon Kids Institute and Joondalup Health Campus.

